# Changes in early aperiodic EEG activity are linked to autism diagnosis and language development in infants with family history of autism

**DOI:** 10.1101/2024.12.15.24319061

**Authors:** Carol L. Wilkinson, Haerin Chung, Amy Dave, Helen Tager-Flusberg, Charles A. Nelson

## Abstract

Delays in language often co-occur among toddlers diagnosed with autism. Despite the high prevalence of language delays, the neurobiology underlying such language challenges remains unclear. Prior research has shown reduced EEG power across multiple frequency bands in 3-to-6-month-old infants with an autistic sibling, followed by accelerated increases in power with age. In this study, we decompose the power spectra into aperiodic (broad band neural firing) and periodic (oscillations) activity to explore possible links between aperiodic changes in the first year of life and later language outcomes. Combining EEG data across two longitudinal studies of infants with and without autistic siblings, we assessed whether infants with an elevated familial likelihood (EFL) exhibit altered changes in both periodic and aperiodic EEG activity at 3 and 12 months of age, compared to those with a low likelihood (LL), and whether developmental change in activity is associated with language development. At 3-months of age (n = LL 59, EFL 57), we observed that EFL infants have significantly lower aperiodic activity from 6.7-55Hz (p<0.05). However, change in aperiodic activity from 3 to 12 months was significantly increased in infants with a later diagnosis of autism, compared to EFL infants without an autism diagnosis (n = LL-NoASD 41, EFL-noASD 16, EFL-ASD 16). In addition, greater increases in aperiodic offset and slope from 3-to12-months were associated with worse language development measured at 18 months (n = 24). Findings suggest that early age-dependent changes in EEG aperiodic power may serve as potential indicators of autism and language development in infants with family history of autism.

**Lay Abstract:** In this study we investigate brain activity in infants with and without a family history of autism in order to understand links between early brain development, autism, and language delays. We find that infants later diagnosed with autism showed altered developmental changes in brain activity during the first year of life, and these changes were associated with poorer language outcomes at 18 months. Findings suggests that early developmental changes in brain activity could help with early identification of infants at increased likelihood for autism and language delays.

## Introduction

Early delays in language have been shown to have long lasting impacts on behavioral functioning, academic success, vocational outcomes and quality of life (Law et al., 2009; Snowling et al., 2006; Young et al., 2002). Language delays are highly prevalent in young children diagnosed with autism. At the time of diagnosis, as many as 40% of autistic toddlers have co-occurring language delays(Reetzke et al., 2022), and the degree of later gains in language development is variable, with 30% of autistic children remaining minimally verbal (Tager-Flusberg & Kasari, 2013). Notably, younger siblings of autistic children are at elevated likelihood for *both* autism and language delays (Marrus et al., 2018). Despite the high prevalence of language impairments in autism, our understanding of the neurobiology underlying impairments is limited. Pinpointing neural markers that are specifically associated with language delays in infant-siblings may shed light on neural mechanisms of early language development and facilitate earlier detection and intervention.

Electroencephalography (EEG) provides a readout of network-level brain activity at the scalp, and has proven to be a promising tool for identifying early neural markers of altered neural development in infancy. In a number of studies resting (baseline) EEG data have been collected longitudinally in infants with an elevated familial likelihood of developing autism (EFL, infants with an older sibling with autism). These studies have observed a global reduction in frontal EEG power across multiple frequency bands at early ages (3-6 months), as well as differences in EEG trajectories from 6 to 24 months in EFL infants regardless of later autism diagnosis (Huberty et al., 2021; Levin et al., 2017; Tierney et al., 2012; Wilkinson et al., 2020). While EEG power is reduced in early infancy in EFL infants, differences are less pronounced at 12 and 24 months, and studies of older autistic children have observed either no differences, or *increases* in absolute gamma power(Arutiunian et al., 2024; Huberty et al., 2021; Mukerji et al., 2024; Rojas & Wilson, 2014; van Diessen et al., 2015). This suggests that *change* in power spectral features early in development may be an important biomarker for autism or other developmental delays. Aligned with this hypothesis, longitudinal EEG power measures collected in the first postnatal year are better predictors of autism outcome than EEG measures collected in the second year, or across the first 3 years of life(Gabard-Durnam et al., 2019).

Studies examining associations between EEG power and language development have had mixed findings depending on age and likelihood group. In infancy, lower frequencies (delta, theta, and alpha) have been associated with language development in both low likelihood infants (LL, infants without an older sibling with autism) and EFL infants(Levin et al., 2017; Pierce et al., 2021). In contrast, in toddlers (16-36 months) frontal gamma power has been associated with language development (Benasich et al., 2008; Gou et al., 2011; Wilkinson et al., 2019). Interestingly, the direction of association between brain activity and language often differs between EFL versus LL toddlers. Gamma power is positively associated with language in LL toddlers, yet negatively associated in EFL toddlers, suggesting possible differences in factors impacting language development between groups (Lombardo et al., 2015; Swanson et al., 2017; Wilkinson et al., 2019). The increased gamma power observed in autistic children has been hypothesized to reflect imbalance in neural excitation/inhibition (E/I)(Brunel & Wang, 2003; Buzsáki & Wang, 2012; Economo & White, 2012). Thus, while higher gamma power in typically developing toddlers and preschoolers may be a beneficial indicator of brain activity for language development, in autistic children it may instead serve as a marker of E/I imbalance impeding processes important for language and cognitive development.

While past EEG studies in autism have largely focused on either absolute or relative measures of power, newer methods utilize parametrization of the spectrum into aperiodic and periodic components to provide a more accurate estimate of non-oscillatory and oscillatory (respectively) activity(Donoghue et al., 2020; Ostlund et al., 2022). The aperiodic signal is defined by the 1/f power law distribution that underlies the absolute power spectra(Gao et al., 2017; Manning et al., 2009; Miller et al., 2009), and can be described by the offset and slope (defined as the χ in the 1/f^χ^ formula). The aperiodic offset is thought to reflect broad band, non-oscillatory neuronal firing(Manning et al., 2009; Miller, 2010), and, growing evidence suggests that the slope of the aperiodic signal may, in part, reflect the excitatory-inhibitory (E/I) balance of the brain, with a flatter slope associated with increased excitation over inhibition(Chini et al., 2022; Gao et al., 2017; McKeon et al., 2024). Together, these components of aperiodic activity may inherently reflect broader neural dynamics such as cortical excitability or developmental maturation. In addition, modulation in aperiodic activity, such as flatter slopes, can influence absolute power in higher frequencies, like gamma. Thus, previous associations between absolute gamma power and language development could instead reflect associations in aperiodic activity and language development. By accounting for individual differences in aperiodic activity, parametrization of the absolute spectrum also provides more accurate measurement of peak frequency and amplitude of oscillatory, or periodic components of the spectra(Buzsáki et al., 2013; Donoghue et al., 2020; Ostlund et al., 2022). Notably, both aperiodic and periodic components change with development, with dynamic changes occurring during the first year after birth(Cellier et al., 2021; Hill et al., 2022; Rayson et al., 2023; Rico-Picó et al., 2023; Wilkinson et al., 2024).

In this study, we aimed to investigate developmental changes over the first postnatal year in aperiodic and periodic components in EFL vs LL infants and assess how alterations may be associated with later autism and language outcomes. Leveraging data collected across two infant-sibling studies with 3– and 12-month time points, we first assessed whether there are differences in frontal aperiodic and periodic power in EEG data collected in LL and EFL infants at 3 and 12 months of age. We focused a priori on frontal regions given prior findings of reduced frontal power in EFL infants and associations between frontal gamma power and language. Next, we determined whether EFL-ASD infants exhibit distinctive patterns of change in EEG aperiodic or periodic power from 3 to 12 months of age. Finally, we assessed whether developmental changes in EEG features are associated with language development.

## Methods

### Participants

This analysis includes infants who were recruited as part of two consecutive studies in the same lab: The Infant Sibling Project and the Infant Screening Project. Both studies were prospective, longitudinal studies, enrolling infants with and without first degree family history of ASD starting as early as 3-months of age. Specifically, the Infant Sibling Project recruited infants at either 3 or 6 months of age and followed them longitudinally at 9, 12, 18, 24, and 36 months. The Infant Screening Project initially recruited infants at 12-14 months of age, but then added a 3-month enrollment option part way through the study.

The Infant Screening Project screened all infants for elevated social communication concerns at 12 months of age based on the Communication and Symbolic Behavior Scale(Wetherby & Prizant, 2012). Given the increased likelihood of both autism and language delay in infants screening positive on the CSBS, these infants were excluded from the LL group in this analysis. Exclusion criteria included gestational age less than 36 weeks, history of prenatal or postnatal medical or neurological problems, as well as identified genetic disorders. Sample characteristics are shown in Table 1. Additional information regarding overall enrollment, visits, and EEG retention based on collection, behavioral exclusions, and data quality exclusions are described in Supplemental Tables 1 and 2.

**Table 1:** Sample characteristics.

|  | 3-month |  | 12-month |  |
| --- | --- | --- | --- | --- |
| Group<br>N (Infant Sibling Project; Infant Screening Project) | LL<br>N = 59 (14, 45) | EFL<br>N = 57 (32, 25) | LL<br>N = 157 (71, 86) | EFL<br>N = 144 (84, 60) |
| **Sex** | 35 M, 22 F | 35 M, 24 F | 82 M, 75 F | 79 M, 65 F |
| **Ethnicity, n (%)** |  |  |  |  |
| Not Hispanic or Latino | 58 (98) | 47 (82) | 150 (96) | 133 (92) |
| Hispanic or Latino | 1 (2) | 9 (16) | 5 (3) | 11 (8) |
| Not Answered | 0 (0) | 1 (2) | 2 (1) | 0 (0) |
| **Race, n (%)** |  |  |  |  |
| White | 50 (85) | 46 (80) | 128 (82) | 129 (90) |
| Black or African American | 1 (2) | 0 (0) | 4 (3) | 1 (1) |
| Asian | 1 (2) | 2 (4) | 5 (3) | 8 (6) |
| Mixed Race | 7 (12) | 8 (14) | 18 (11) | 6 (4) |
| Not answered | 0 (0) | 1 (2) | 2 (1) | 0 (0) |
| **Household income, % (n)\*** |  |  |  |  |
| <\$35,000 | 1 (2) | 4 (7) | 6 (4) | 6 (4) |
| \$35,000 - \$75,000 | 2 (3) | 3 (5) | 7 (4) | 15 (10) |
| >\$75,000 | 54 (92) | 43 (75) | 130 (83) | 103 (72) |
| Not answered | 2 (3) | 7 (12) | 14 (9) | 20 (14) |
| **EEG quality metrics** |  |  |  |  |
| Number of Segments | 102.6 ± 38.7 | 96.7 ± 35.1 | 91.3 ± 40.5 | 93.1 ± 52.8 |
| Percent Good Channels | 92.5 ± 4.7 | 92.7 ± 4.9 | 92.3 ± 4.4 | 92.3 ± 4.6 |
| Percent ICs Rejected | 0.33 ± 0.10 | 0.36 ± 0.13 | 0.39 ± 0.10 | 0.37 ± 0.12 |
| Mean Artifact Probability of Kept ICs. | 0.15 ± 0.05 | 0.15 ± 0.04 | 0.12 ± 0.05 | 0.12 ± 0.05 |
| **# of participants with 3 & 12 month EEG data**<br>N (Infant Sibling Project; Infant Screening Project) | N = 47 (12, 35) | N = 36 (21, 15) | N = 47 (12, 35) | N = 36 (21, 15) |

**Abbreviations:** LL: Low Likelihood; EFL: Elevated Likelihood; M: Male; F: Female; IC: Independent Components. \*One 3-month infant, and Four 12-month infants reported to have income of \$30-39,000 was placed in <\$35,000 category. Three 12-months infants with reported income of \$70-79,000 placed in >\$75,000 category.

Institutional review board (IRB) approval was obtained from Boston Children’s Hospital (The Infant Sibling Project (IRB-X06-08-0374), and the Infant Screening Project (IRB-P00018377). Secondary analysis for this paper was also approved (IRB-P00037531). Written informed consent was obtained from a parent or guardian prior to each child’s participation in the study.

### Developmental assessments

Participants in both studies were seen longitudinally for developmental assessments using the Mullen Scales of Early Learning (MSEL). As the Infant Screening Project spanned the COVID-19 pandemic, in-person assessments for a number of participants at 24 and 36 months was not possible and remote evaluations took place. Therefore, exploratory analyses related to language development utilized the MSEL Verbal Developmental Quotient (VDQ) at 18 months when more in person visits were completed in order to have a sufficiently power sample to assess brain-language associations. The Verbal Developmental Quotient is calculated by averaging age-equivalents for receptive language and expressive language scales, dividing by the child’s chronological age, and multiplying by 100. We acknowledge that language assessments at 18 months are less stable, and that as many as 60% of toddlers with delays at 18-24 months will “catch up” by preschool(Paul, 1993). However, late talkers regardless of persistence of delays, are at elevated risk for experiencing later academic and social-emotional challenges, and thus identifying early predictors of 18 month language development is also clinically valuable.

### Autism assessments

For the Infant Sibling Project, ASD outcomes were determined using the Autism Diagnostic Observation Schedule(Lord & Rutter, 2012) (ADOS) and parent-child interaction administered at 24 and/or 36 months of age. During the COVID-19 pandemic, remote autism evaluations for the Infant Screening Project included the Brief Observation of Symptoms of Autism(Dow et al., 2022) (BOSA), parent-child interaction, and Vineland Adaptive Behavioral Scales – Third Edition, Parent Interview Form(Sparrow, 2018) (Vineland-3). High convergent validity between the BOSA and the ADOS-2 has been demonstrated(Dow et al., 2022). In both studies, for toddlers meeting criteria on the ADOS/BOSA or coming within three points of cutoffs, a licensed clinical psychologist reviewed scores and video recordings of concurrent and previous behavioral assessments. A best estimate of clinical judgement was determined based on DSM-5 criteria for autism diagnosis. Of the 179 EFL participants who completed outcome visits, 31% (57/179) met clinical judgement for autism.

### EEG data collection

In both studies, “resting”, non-task-related, EEG data were collected while the infant was held by their seated caregiver in a dimly lit, sound attenuated room with low-electrical signal background. In the Infant Sibling Study, a research assistant ensured the infant remained calm by blowing bubbles and/or showing toys. Research assistants refrained for social interactions with infants. Continuous EEG data was collected using 64-channel Geodesic Sensor (∼10% of data in analysis) or a 128-channel Hydrocel Geodesic Sensor Nets (Electrical Geodesics, Inc., Eugene, OR), connected to either a NetAmps 200 or 300 amplifier (Electrical Geodesic Inc.) and sampled at either 250 or 500Hz. The switch in equipment occurred because the company ceased supporting the 64-channel net equipment during the longitudinal study. See Supplemental Tables 3 and 4 for breakdown of EEG collection across nets and amplifiers. Possible differences in data quality or EEG measures based on NetAmp 200 vs NetAmp 300 collection parameters was assessed and no significant differences were observed (Supplemental Table 5).

In the Infant Screening Project, a video of abstract moving objects was shown and EEG data was collected with the 128-channel Hydrocel Geodesic Sensor Net, connected to a NetAmps 300 amplifier and sampled at 500Hz. For both studies, data were referenced online to a single vertex electrode (Cz). Electrooculographic electrodes were removed to improve the child’s comfort.

## EEG pre-processing and rejection criteria

Raw Netstation (Electrical Geodesics, Inc) files were exported to MATLAB (version R2017a) for preprocessing and absolute power calculations using the Batch Automated Processing Platform (BEAPP(Levin et al., 2018)) with integrated Harvard Automated Preprocessing Pipeline for EEG (HAPPE(Gabard-Durnam et al., 2018)). For each EEG, a 1Hz high-pass and 100Hz low-pass filter were applied, data sampled at 500Hz were resampled to 250Hz, and then run through the HAPPE module consisting of 60Hz line noise removal, bad channel rejection, and artifact removal using combined wavelet-enhanced independent component analysis (ICA) and Multiple Artifact Rejection Algorithm (MARA^5,6^). As two net types were used, channels for ICA with MARA were selected from spatial locations that corresponded across nets (EGI, Eugene, OR) The following channels, in addition to the 10-20 electrodes, were used for MARA: 64-channel net – 3, 8, 9, 16, 18, 21, 25, 30, 32, 33, 38, 41, 43, 45, 50, 53, 57, 58; and 128-channel net – 4, 13, 19, 28, 37, 41, 47, 55, 65, 67, 75, 77, 87, 90, 98, 103, 112, 117. See Supplemental Figure1 for mapping of both nets. After artifact removal, channels removed during bad channel rejection were then interpolated, data were referenced to the average reference, detrended to the signal mean, and segmented into 2-second segments. Any segments with retained artifact were rejected using HAPPE’s amplitude and joint probability criteria. EEG recordings were rejected using the following HAPPE data quality measures: Fewer than 20 segments (40 seconds of total EEG), percent good channels < 80%, percent independent components rejected >80%, mean artifact probability of components kept > 0.3, and percent variance retained < 25%. Table 1 shows quality metrics across LL and EFL groups. There were no significant differences between groups.

## EEG Power Spectra Analysis

For each 2-second segment, the power spectral density at each electrode was calculated in the BEAPP Power Spectral Density (PSD) module using a multitaper spectral analysis(Babadi & Brown, 2014) and three orthogonal tapers. For each electrode, the PSD was averaged across segments, and then averaged across all frontal electrodes [64-channel: 8 (F1), 3 (F2), 3 (F3), 62 (F4), 16 (FC5), 57 (FC6); 128-channel net: 19 (F1), 4 (F2), 24 (F3), 124 (F4), 28 (FC5), 117 (FC6), 11 (FZ)]. The PSD was then further analyzed using a modified version of SpecParam v1.0.0(Donoghue et al., 2020) (https://github.com/fooof-tools/fooof; in Python v3.6.8; modifications described in (Wilkinson et al., 2024) with available code at osf.io/u3gp4. The SpecParam model was used in the fixed mode (no spectral knee) evaluating spectra between 2-55Hz, with *peak_width_limits* set to [0.5, 18.0], *max_n_peaks* = 7, and *peak_threshold* = 2). Mean R^2^ for the full sample using this modified version of SpecParam was 0.997 (STD 0.009; range 0.875-0.9998). Mean R^2^ for EFL and LL groups and age bins ranged from 0.992-0.999. No EEGs were rejected due to poor fit by SpecParam.

Using Specparam with a fixed mode, the aperiodic component is modeled as L = b – log(Fx), where b is the offset, χ is the exponent, and *F* is a vector of input frequencies. This function is the same as fitting a line in log-log space, where the offset in the equation above is the y-intercept. As previously described in Wilkinson et al., 2024, here we define aperiodic offset as the intercept at 2.5Hz (rather than 0Hz), as there are high levels of error in the SpecParam estimates at frequencies below 2.5Hz(Wilkinson et al., 2024). Aperiodic slope, defined by χ in the above equation is provided by SpecParam. Total aperiodic activity is defined as the integral of the aperiodic spectra from 2.5-55Hz. To characterize the periodic power spectra, the SpecParam estimated aperiodic signal was subtracted from the absolute power spectrum. Periodic gamma power was calculated using the integral of the periodic spectra from 30-45Hz.

## Statistical Analyses

Group differences in the absolute, aperiodic, and periodic power spectra (averaged across frontal electrodes) were assessed using a non-parametric clustering method controlling for multiple comparisons using Monte Carlo estimation with 10,000 permutations(Maris & Oostenveld, 2007), employed with the MNE-Python(Gramfort et al., 2014) ‘permutation_cluster_test’ function. This method is a data driven approach to identifying frequency clusters that are significantly different between groups, while also controlling for multiple comparisons. Group differences in EEG measures were statistically compared using Mann Whitney U (two-sided) for likelihood group comparisons, or Kruskall Wallis for outcome group comparisons, using the Scipy functions in Python (version 3.7.6).

Mixed-effects linear regression models were performed to test whether change in aperiodic features (slope, offset, total aperiodic activity) were different between outcome groups (LL-NoASD, EFL-NoASD, EFL-ASD). Age (months) was treated as a within-subjects factor and outcome group were included as between-subjects factor. Sex was included as a covariate in analyses as there were differences in sex distribution between outcome groups (with the EFL-ASD group having higher proportion of males). Models were fit using lmer4 package (version 1.1-35.5) and figures were generated using the emmip function from emmeans package (version 1.10.5) in R (version 4.3.2).

To test associations between change in EEG measures and language skills, linear regressions were performed with 18-month verbal developmental quotients as the dependent variable, developmental change from 3-to 12-months in aperiodic measures as the independent variable (Model 1), and either 18m nonverbal developmental quotients (Model 2) or ASD diagnosis included as a covariate (Model 3). Models were fit using the statsmodel ols function in Python (version 3.7.6).

Study data were collected and managed using REDCap electronic data capture tools hosted at Boston Children’s Hospital(Harris et al., 2009, 2019).

## Results

Table 1 shows sample characteristics for participants providing EEG data either at 3-months or 12-months of age. Fewer EEGs were available at 3 months of age for both studies, as the early time point was not included until part-way through each study. Forty-one LL-NoASD, 16 EFL-ASD, and 16 EFL-NoASD infants had EEG data at both 3 and 12 months.

### Power spectra differences at 3 months of age

Frontal absolute, aperiodic, and periodic power spectra for LL and EFL infants at 3 months are shown (Figure 1A). We used a non-parametric clustering method, controlling for multiple comparisons, to assess for group differences in the power spectra. Within the 3-month absolute power spectra a marginally significant cluster was identified in the gamma range (41.4-55Hz, p=0.05). Within the 3-month aperiodic component, a significant cluster spanning alpha through gamma frequencies was identified (6.8-55Hz, p < 0.05). No significant clusters were identified in the 3-month periodic spectra. The combination of these findings suggest that differences observed in absolute gamma power are largely driven by differences in aperiodic activity. Statistically significant group differences were observed for both absolute gamma power and total aperiodic activity, with the EFL group exhibiting reduced activity (Mann Whitney U (EFL vs LL); Absolute Gamma: U = 1268, p < 0.05, adj-p <0.1; Total Aperiodic Activity: U = 1187, p < 0.01, adj-p <0.05; Figure 1B, C, D) compared to the LL group. No significant difference was observed for periodic gamma power, aperiodic slope or offset. Given differences in the nets and amplifier used during data collection, sensitivity analyses we performed. Similar findings were observed when removing EEGs either recorded with 64-channel nets or with a NetAmp 200 Amplifier for the 3-month time point (Supplemental Table 7) and similarly, no significant differences were observed at 12 months.

**Figure 1.**
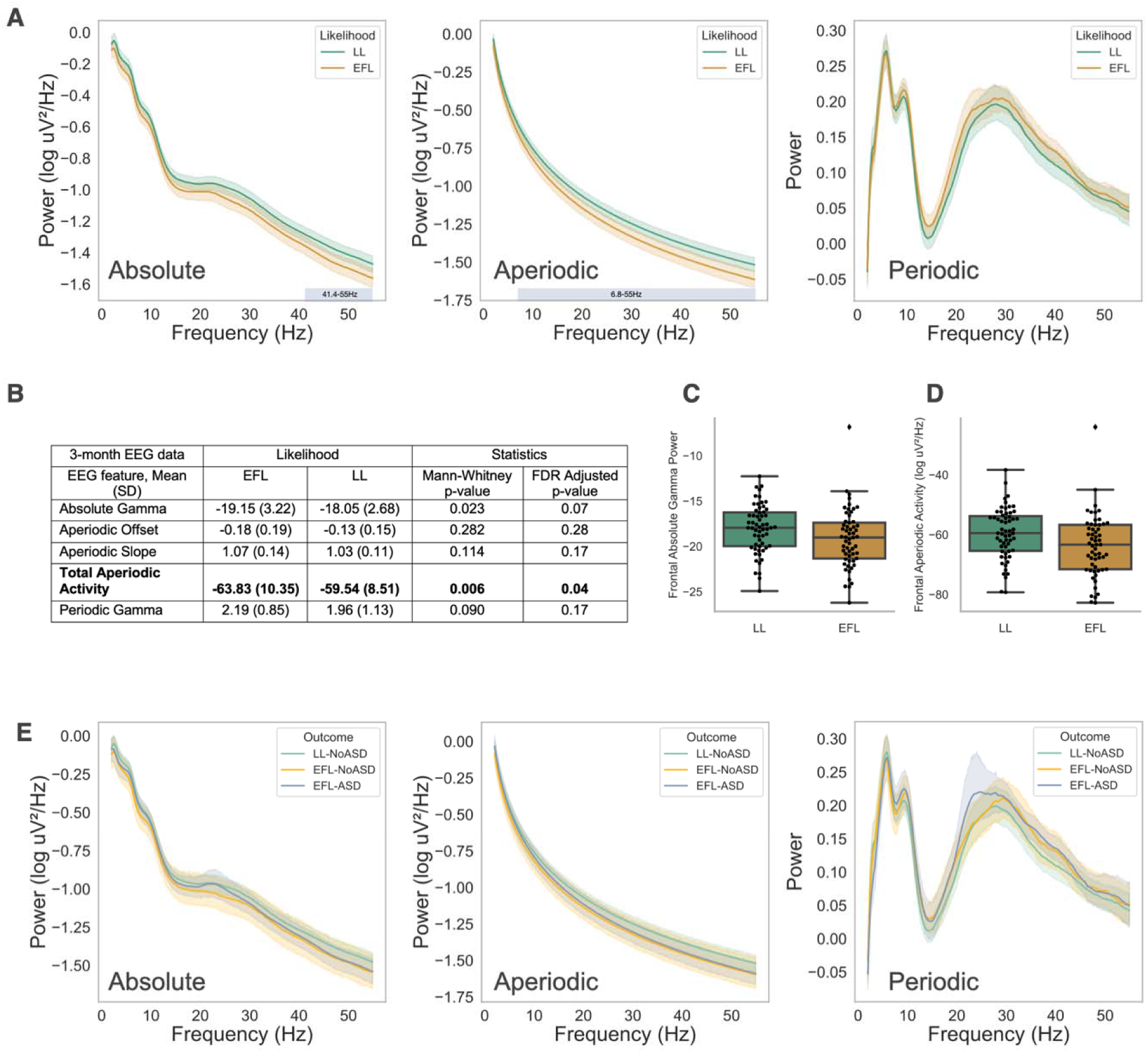
(**A**) Absolute, Aperiodic, and Periodic spectra at 3 months by Likelihood group. Colored shading around spectra represents 95% confidence intervals. Identified statistically significant clusters are shown as horizontal bars defined by the cluster’s frequency band. (**B**) Mean comparison of EEG measures by Likelihood group using two-sided Mann Whitney U test. (**C, D**) Box plot of group differences in absolute gamma power and total aperiodic activity. (**E**) Absolute, aperiodic, and periodic spectra at 3 months by Outcome group. Shading around spectra represents 95% confidence intervals.

Next, we assessed whether differences in aperiodic activity at 3 months were driven by those infants later diagnosed with autism. Power spectra from EEG collected at 3 months of age are shown in Figure 1E based on later autism outcome (LL-NoASD n = 47, EFL-NoASD n = 22, EFL-ASD n = 21). No significant differences were observed between outcome groups on non-parametric clustering methods across the spectrum and no significant differences were observed for specific aperiodic or gamma power measures (Kruskal Wallis Test, p-value range 0.13-0.66; Supplemental Table 4)

### Power spectra differences at 12 months

Power spectra for LL and EFL infants at 12 months are shown in Figure 2A, and 12-month spectra group by later autism outcomes are shown in Figure 2B (LL-noASD n = 143, EFL-NoASD n = 85, EFL-ASD n = 41). Unlike at 3 months, there were no significant differences between groups on any measures.

**Figure 2.**
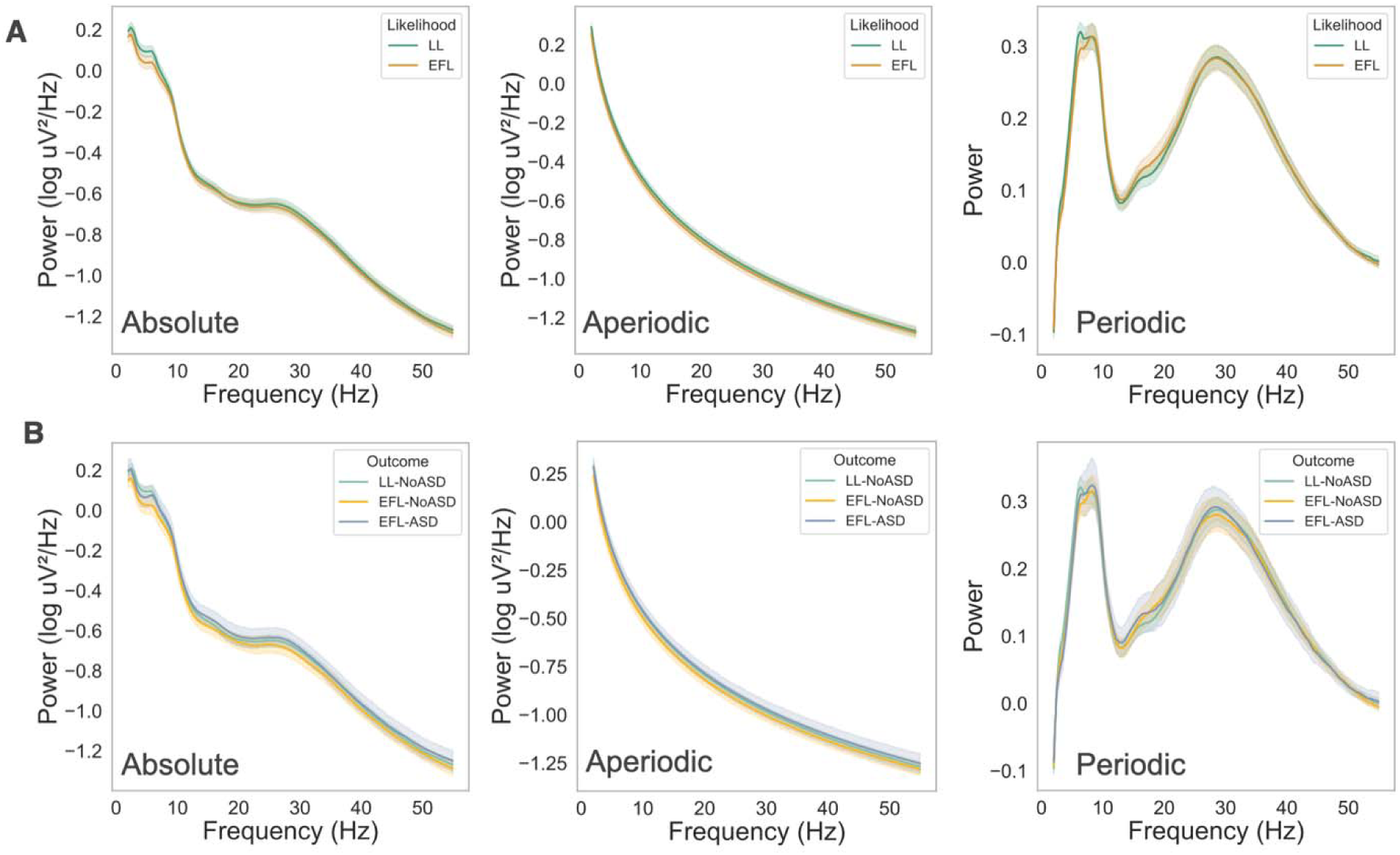
Absolute, Aperiodic, and Periodic spectra at 12 months groups by (A) Likelihood and (B) Outcome groups. Shading represent 95% confidence intervals.

### Change in EEG aperiodic and periodic activity from 3 to 12 months

Our 3– and 12-month-old findings suggest that EFL infants with significantly lower aperiodic activity early in infancy have greater increases aperiodic activity between 3 and 12 months, leading to similar EEG aperiodic spectra at 12 months. This finding might reflect greater developmental increases in aperiodic activity exhibited by the EFL group, regardless of ASD outcome. Alternatively, it could be primarily driven by more pronounced changes in aperiodic activity among the EFL-ASD infants. To test this, we limited our analysis to those infants with both 3– and 12-month-old data, and confirmed that outcome groups were similar in the number of days between visits. 41 LL-NoASD, 16 EFL-ASD, and 16 EFL-NoASD infants had EEG data at both 3 and 12 months.

A priori pairwise comparisons revealed significant outcome group differences in the change in EEG measures from 3 and 12 months (Table 2 and Figure 3; see Supplementary Table 9 for full model outputs). Specifically, EFL-ASD infants exhibited greater change in aperiodic total activity when compared to LL-NoASD infants, and greater change in aperiodic offset when compared to both EFL-NoASD and LL-NoASD infants. However, differences in offset between EFL-ASD and LL-NoASD infants remained significant after correcting for multiple comparisons. Given prior literature on absolute gamma activity in autism, we also tested whether outcome groups differed in change of absolute gamma power. No significant differences were found, although similar patterns were observed (Table 2 and Figure 3).

**Figure 3.**
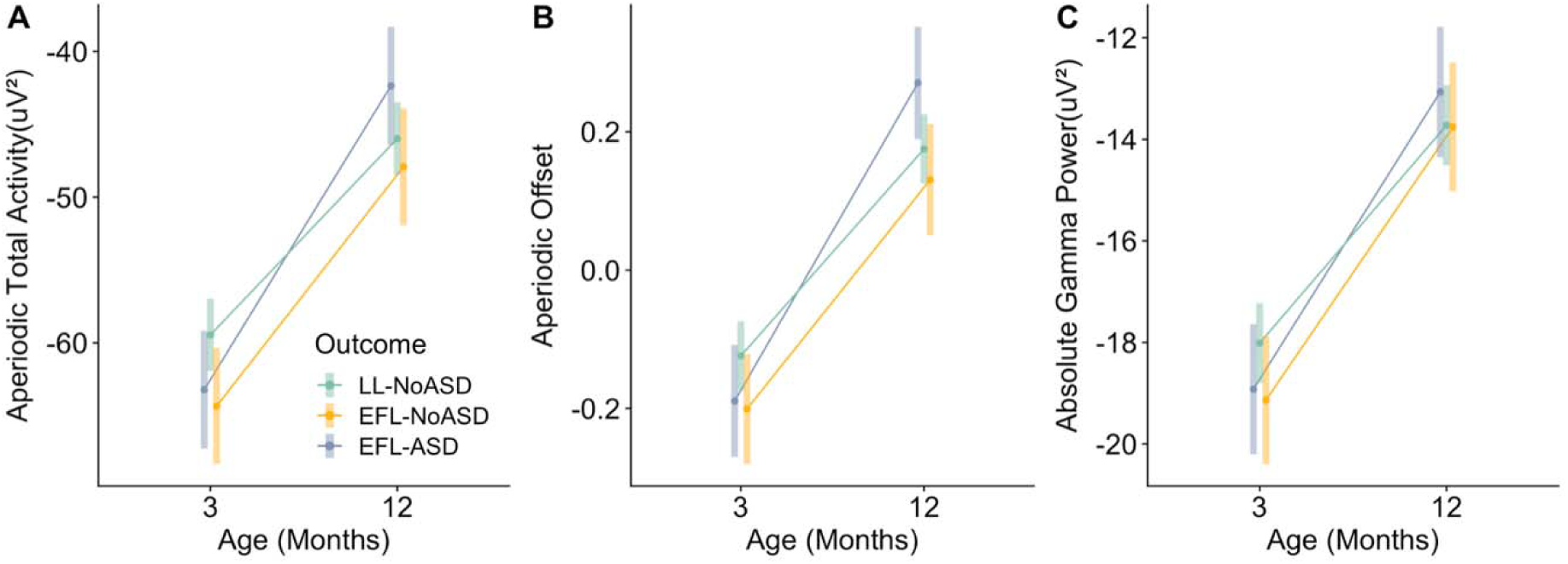
Change in EEG measures from 3 to 12 months. (A) Aperiodic Total Activity, (B) Aperiodic Offset, and (C) Absolute Gamma Power.

**Table 2:** A priori pairwise comparisons of EEG changes between outcome groups (3-to-12 months)

| EEG Measure | Outcome Comparison | estimate | SE | t-statistic | p-value | p-value adjusted |
| --- | --- | --- | --- | --- | --- | --- |
| Aperiodic Total Activity | (EFL-ASD) - (LL-NoASD) | -7.39 | 3.23 | -2.29 | 0.03 | 0.08 |
|  | (EFL-ASD) - (EFL-NoASD) | -4.43 | 3.83 | -1.15 | 0.25 | 0.36 |
|  | (LL-NoASD) - (EFL-NoASD) | 2.96 | 3.23 | 0.92 | 0.36 | 0.36 |
| Aperiodic Offset | (EFL-ASD) - (LL-NoASD) | -0.16 | 0.06 | -2.55 | 0.01 | 0.04 |
|  | (EFL-ASD) - (EFL-NoASD) | -0.13 | 0.07 | -1.72 | 0.09 | 0.14 |
|  | (LL-NoASD) - (EFL-NoASD) | 0.03 | 0.06 | 0.51 | 0.61 | 0.61 |
| Aperiodic Exponent | (EFL-ASD) - (LL-NoASD) | -0.02 | 0.04 | -0.55 | 0.59 | 0.59 |
|  | (EFL-ASD) - (EFL-NoASD) | -0.05 | 0.05 | -1.01 | 0.32 | 0.59 |
|  | (LL-NoASD) - (EFL-NoASD) | -0.03 | 0.04 | -0.65 | 0.52 | 0.59 |
| Absolute Gamma Power | (EFL-ASD) - (LL-NoASD) | -1.56 | 0.95 | -1.64 | 0.1 | 0.31 |
|  | (EFL-ASD) - (EFL-NoASD) | -0.47 | 1.13 | -0.42 | 0.68 | 0.68 |
|  | (LL-NoASD) - (EFL-NoASD) | 1.09 | 0.95 | 1.15 | 0.26 | 0.38 |

### Exploratory analysis of EEG features associated with language development

Language and cognitive impairments often co-occur in autistic toddlers, and therefore EEG findings associated with autism outcomes may not be specific to the diagnosis, but rather reflective of developmental delays. In addition, while we have previously found that increased absolute gamma power measured at both 18 and 24 months is associated with reduced language scores in EFL infants, our current analyses, now parametrizing the power spectra into aperiodic and periodic components, suggest that associations between absolute gamma power and language in autism may instead be driven by alterations in underlying aperiodic activity. Given our above observation that increased change in aperiodic offset and total aperiodic activity are associated later ASD diagnosis, we next explored whether change in aperiodic activity is associated with future language impairment across the EFL group (n = 24) when controlling for either autism outcome or nonverbal cognitive ability. Linear regressions were performed with 18-month verbal developmental quotients as the dependent variable, developmental change from 3-to 12-months in aperiodic measures as the independent variable (Model 1), and either 18m nonverbal developmental quotients (Model 2) or ASD diagnosis included as a covariate (Model 3). Results are shown in Table 3. Increased changes in aperiodic offset and slope were significantly associated with reduced 18-month verbal developmental quotients (Figure 4). Notably, associations remained significant when accounting for differences in 18-month nonverbal developmental quotient or for autism diagnosis. No significant associations were observed for aperiodic total activity or absolute gamma power.

**Figure 4.**
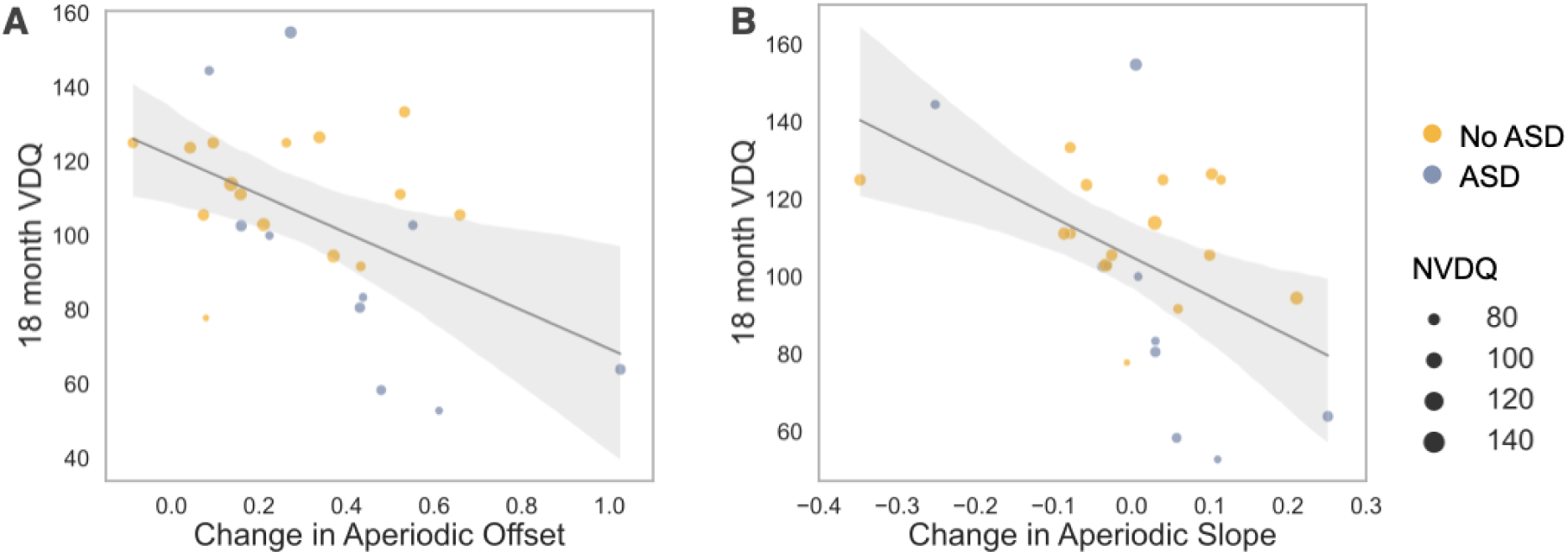
Regression plots from Model 1 comparing (A) change in aperiodic offset and 18-month MSEL verbal developmental quotient (VDQ) and (B) change in aperiodic slope and 18-month MSEL VDQ. Size of dots indicated individual’s MSEL nonverbal developmental quotient (NVDQ). Color represents autism outcome.

**Table 3:**
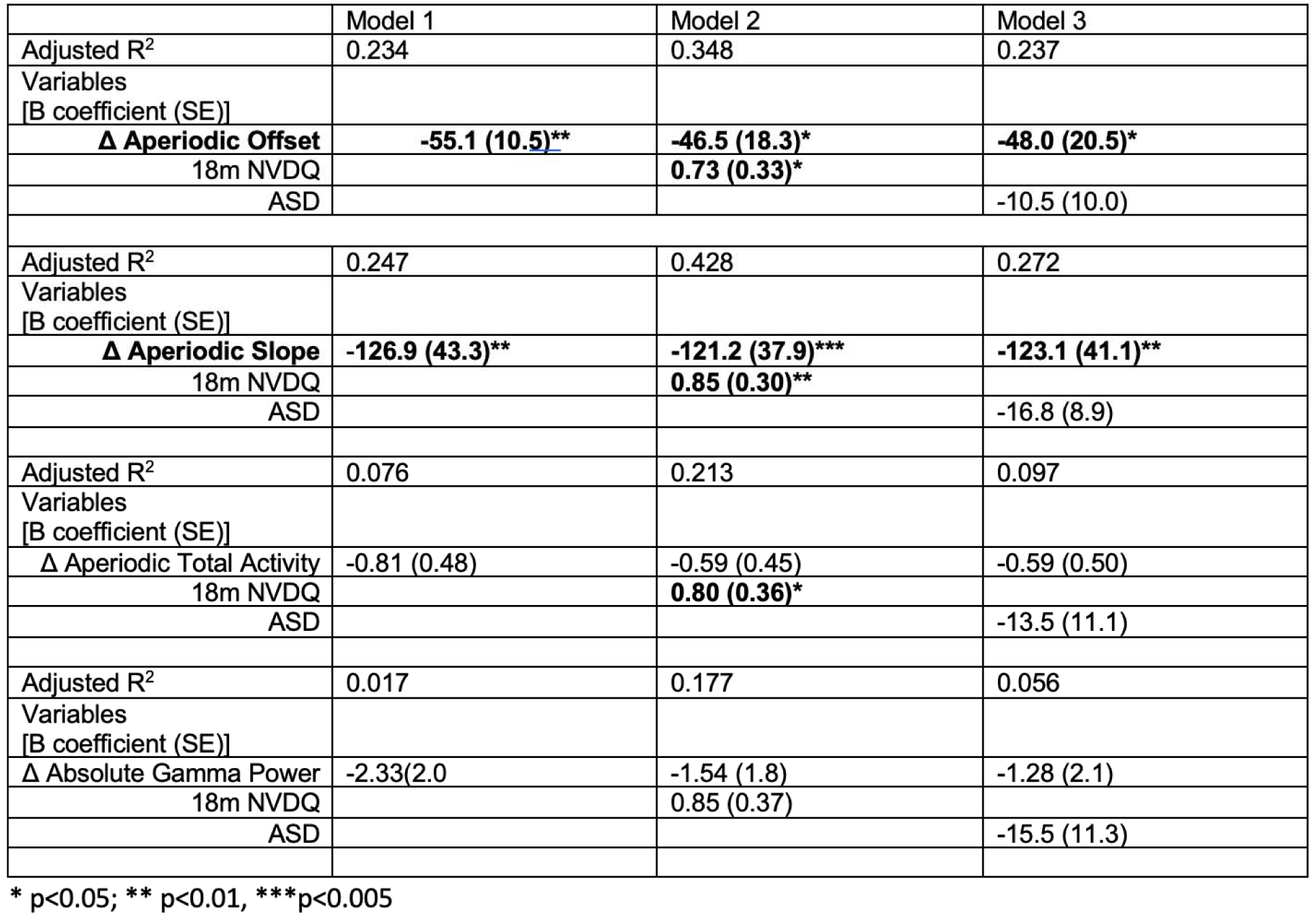
Linear regressions assessing associations between developmental change EEG features and 18-month MSEL VDQ.

|  | Model 1 | Model 2 | Model 3 |
| --- | --- | --- | --- |
| Adjusted R<sup>2</sup> | 0.234 | 0.348 | 0.237 |
| Variables<br>[B coefficient (SE)] |  |  |  |
| **Δ Aperiodic Offset** | **-55.1 (10.5)\*\*** | **-46.5 (18.3)\*** | **-48.0 (20.5)\*** |
| 18m NVDQ |  | **0.73 (0.33)\*** |  |
| ASD |  |  | -10.5 (10.0) |
| Adjusted R<sup>2</sup> | 0.247 | 0.428 | 0.272 |
| Variables<br>[B coefficient (SE)] |  |  |  |
| **Δ Aperiodic Slope** | **-126.9 (43.3)\*\*** | **-121.2 (37.9)\*\*\*** | **-123.1 (41.1)\*\*** |
| 18m NVDQ |  | **0.85 (0.30)\*\*** |  |
| ASD |  |  | -16.8 (8.9) |
| Adjusted R<sup>2</sup> | 0.076 | 0.213 | 0.097 |
| Variables<br>[B coefficient (SE)] |  |  |  |
| Δ Aperiodic Total Activity | -0.81 (0.48) | -0.59 (0.45) | -0.59 (0.50) |
| 18m NVDQ |  | **0.80 (0.36)\*** |  |
| ASD |  |  | -13.5 (11.1) |
| Adjusted R<sup>2</sup> | 0.017 | 0.177 | 0.056 |
| Variables<br>[B coefficient (SE)] |  |  |  |
| Δ Absolute Gamma Power | -2.33(2.0) | -1.54 (1.8) | -1.28 (2.1) |
| 18m NVDQ |  | 0.85 (0.37) |  |
| ASD |  |  | -15.5 (11.3) |

\* p<0.05; \*\* p<0.01, \*\*\*p<0.005

## Discussion

Leveraging two longitudinal data sets with a prospective infant-sibling design, we compared early developmental changes in EEG power spectra from 3-to 12-months between infants with low and elevated likelihood based on family history of autism. To do this, we both used traditional assessment of absolute EEG power, but also separately analyzed aperiodic and periodic activity. We observe two major findings: First we observe early differences in aperiodic activity in infants with family history of autism regardless of later autism outcome. Second, we observe that greater developmental increases in aperiodic activity are associated with both later autism diagnosis and reduced language development in EFL infants.

### Early differences in EEG activity at 3 months associated with autism family history

At 3-months of age, EFL infants exhibited significantly reduced absolute gamma power as well as overall reduced aperiodic activity compared to LL infants. Interestingly, these differences were not driven by those infants with later diagnoses of autism, suggesting that while early differences in EEG activity may be driven by genetic/familial factors associated with autism, reduced 3-month aperiodic activity alone is not predictive of later diagnosis. We, and others have previously reported reduced EEG power in EFL infants early in infancy(Huberty et al., 2021; Levin et al., 2017; Tierney et al., 2012; Wilkinson et al., 2020), but until now, studies have not examined whether this is due differences in aperiodic or periodic activity. Our observations suggest that prior reported differences in absolute power, could be largely driven by reduced aperiodic activity in EFL infants.

Aperiodic activity represents non-oscillatory, broad band neuronal spiking activity(Manning et al., 2009). The most dramatic increase in aperiodic activity occurs in the first months after birth and likely reflects increases in neuronal number, neuronal connections and synaptogenesis(Wilkinson et al., 2024). In addition, growing evidence suggests that aperiodic slope is an indirect measurement of network excitatory and inhibitory (E/I) balance(Chini et al., 2022; Gao et al., 2017; McKeon et al., 2024). While it is hypothesized that alterations in E/I balance play a role in the development of autism and other neurodevelopmental disorders, we did not observe significant differences in aperiodic slope based on familial likelihood or later outcomes. Thus, reductions in aperiodic activity observed in EFL infants may reflect early hypoconnectivity and/or delays in synaptogenesis.

### Change in aperiodic EEG activity associated with later autism diagnosis

Consistent with previous studies of absolute EEG power, at 12-months, we observed no significant group differences (likelihood or outcome group) for either aperiodic or periodic activity. However, group differences were observed when assessing the *change* in aperiodic activity from 3-to 12-months of age. Here we found infants with later ASD diagnoses exhibited *greater* developmental increases in aperiodic total power and offset. These findings are aligned with longitudinal MRI studies of infant-sibling design which find that EFL-ASD infants exhibit increased growth rates in cortical surface area during the first year of life and increased total brain volume growth rates in the second year compared to both EFL-NoASD and LL infants(Hazlett et al., 2017).

Again, changes in aperiodic slope were not different between likelihood or outcome groups. However, it is important to note that these findings do not directly conflict with prior literature finding altered E/I balance in children or adults with autism. Our observations are specific to EEG activity across the first year after birth, when networks are being established and some of the most significant changes in aperiodic and periodic activity occur. It is still unclear whether aperiodic slope measured during the first year of life is an accurate proxy for E/I balance.

### Greater early developmental change in aperiodic activity associated with reduced language development

Finally, exploratory analysis found that for EFL infants, increased change in aperiodic offset and slope from 3-to-12months was associated with lower language ability at 18 months. Importantly this association remained significant after accounting for either differences in nonverbal development or autism diagnosis.

This is the first study to examine change in EEG activity between these specific ages. Levin et al 2017, focusing on 3-month EEG data from the Infant Sibling Project (41% 3-month EEG overlap with this analysis) showed that increased high alpha absolute power at 3 months was associated with better 12-month expressive language skills. However, Levin et al. 2017 did not parametrize data into aperiodic and periodic components. Huberty et al. (2023), specifically investigated whether change in absolute EEG spectral power from 6 to 36 months is associated with language development in infant-siblings. In Huberty et al., significant associations were limited to 6-month alpha power and *concurrent* language ability, with increased 6-month alpha power associated with better expressive language skills at 6-months, while controlling for concurrent nonverbal skills. In contrast to our findings, Huberty et al found no significant associations between change in EEG power from 6 to 36 months and later language abilities. Differences in both the age range analyzed, and EEG processing methods (absolute vs aperiodic activity) may explain the difference in findings.

A recent study by Piazza et al 2023(Piazza et al., 2023), evaluated EEG data collected at 6 and 12 months of age in infants with either elevated likelihood for autism or language impairment. They observed reduced absolute power in low frequency bands (delta, theta, low alpha) at both ages in infants with elevated autism likelihood compared to those with low autism likelihood. Evaluation based on later outcomes observed that infants with later autism diagnosis also showed reduced delta and theta power at 6 months, and in theta at 12 months, and infants with later language delays showed increased high frequency power at 12 months of age. However, authors did not specifically assess *within subject change* in EEG power, and it is possible that the observed differences at 6 and 12 months, reflect greater changes in aperiodic activity in those infants with later autism diagnoses.

Why might greater changes in aperiodic offset and slope be associated with reduced language development in infants with family history of autism? EFL infants as a group also exhibited significantly lower aperiodic activity at 3 months compared to LL infants. While it is still unclear what neurobiological mechanisms underly this reduced activity, it is possible that EFL infants who overcompensate for these early differences have altered processing of sensory information critical for language development. For example, infants undergoing a rapid shift in connectivity or synaptogenesis in response to underlying aberrant circuitry may not make appropriate connections in response to the sensory environment. In addition, early alterations in aperiodic activity maybe impact the development of neural circuits important for rhythmic activity that play a critical role in language processing and therefore language acquisition.

We also note that for both LL and EFL infants, aperiodic activity increases from 3-to12-months of age. This contrasts prior developmental studies spanning the preschool to adolescent period, where aperiodic activity (both aperiodic offset and slope) is observed to decrease with age(Cellier et al., 2021; Hill et al., 2022; McSweeney et al., 2021; Schaworonkow & Voytek, 2021). Indeed, reduced aperiodic offset and reduced (shallower) slope is often referenced as indicative of more advanced maturation. However, our findings during the first year suggest a different process impacting aperiodic activity during early brain maturation; during this early period we hypothesize that increases in aperiodic activity likely reflect dramatic increases in neuronal number, synaptogenesis, and myelination occurring the during the first year. Whereas in later developmental stages reductions in aperiodic activity may reflect synaptic pruning and refining of networks. Rico-Pico et al. 2023(Rico-Picó et al., 2023) have reported similarly increases in aperiodic offset from 6 to 16 months of age, and Wilkinson et al. 2025 recently reported non-linear changes in aperiodic offset, with the greatest changes between 2-and 12-months of age, followed by a plateau from 12-36 months. Longitudinal studies from birth to school age will be important in determining the inflection point from increases to decreases in aperiodic activity. In this vein, McSweeny et al 2023 (McSweeney et al., 2023)report increases in aperiodic offset and exponent from 4 to 7 years of age, followed by decreases in both components through 11 years of age.

### Limitations

While we combined data from two longitudinal studies, increasing our 3– and 12-month samples sizes, only 73 infants provided data at both time points. Larger prospective longitudinal studies starting early in infancy are required to replicate our findings. In addition, the COVID-19 pandemic limited in person assessments at 24 and 36 months of age, and we were unable to have a sufficient sample size to assess EEG associations with later language development. It is therefore still unknown whether changes in EEG aperiodic activity are predictive of persistent language delay past 18 months of age. Our sample was also largely comprised of infants from White families with higher levels of income, reducing generalizability. We have previously observed that lower household income is also associated with reduced EEG absolute power at 2-3 months of age, with greater increases in power from 3 to 9 months(Wilkinson et al., 2023). Larger datasets with greater variability in SES measures are needed to test whether SES factors such as income, parental education, and stress, moderate our observed associations between EEG aperiodic activity and autism or language outcomes.

### Conclusions

Findings presented in this paper suggest that a combination of early differences in brain development and possible overcompensation for these differences is associated with autism and delayed language development. Future prospective EEG research in infants with increased likelihood should consider prioritizing longitudinal data collection in the first year, including during the first months after birth if possible. Given the close association between ASD and language development, larger longitudinal studies that include EEG data in early infancy are also needed to further tease apart the complex interactions between brain and behavior associations.

## Data Availability

Consents obtained from human participants at our institution for the studies described prohibit sharing of identifiable and de-identified individual data without a data use agreement in place. Please contact the corresponding author with data requests.

## Acknowledgements

We thank all the children and families who generously participated in this research. We thank all the research staff involved in participant recruitment, data collection, and database administration. *Funding Statement*: This research was supported by the National Institutes of Health (R01-DC010290 to C.A.N. and HTF, K23DC07983 and T32MH112510 to C.L.W.)

## Author Contributions

Carol Wilkinson: conceptualization, methodology, software, formal analysis, data curation, writing – original draft, and visualization. Haerin Chung: methodology, software, formal analysis, data curation, writing – original draft, and visualization. Amy Dave: formal analysis, data curation, writing – review & editing. Helen Tager-Flusberg: investigation, resources, writing – review & editing, supervision, project administration, and funding acquisition. Charles A. Nelson: investigation, resources, writing – review & editing, supervision, project administration, and funding acquisition.

