## Supplemental Materials for "Changes in early aperiodic EEG activity are linked to autism diagnosis and language development in infants with family history of autism"

**Supplemental Figure 1 - Electrode layout:** (A) 128-channel Hydrocel Geodesic Sensor Net. (B) 64-channel Geodesic Sensor Net. Pink circles denote 10-20 electrodes, and blue circles denote the additional electrodes included in ICA and MARA steps of pre-processing.

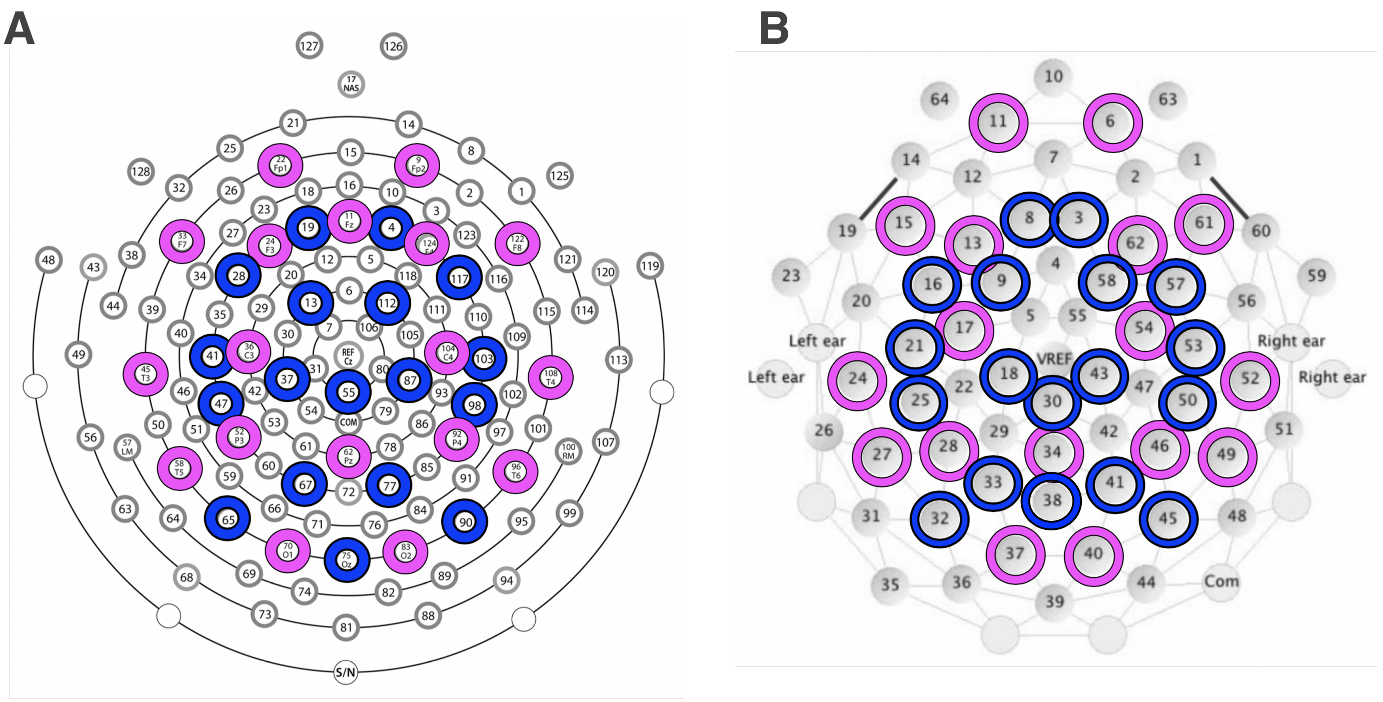

**Supplemental Figure 2:** Here we show comparison of error in SpecParam model estimates by Likelihood or Outcome group. On the right, the estimated Aperiodic Fit (blue) is shown.

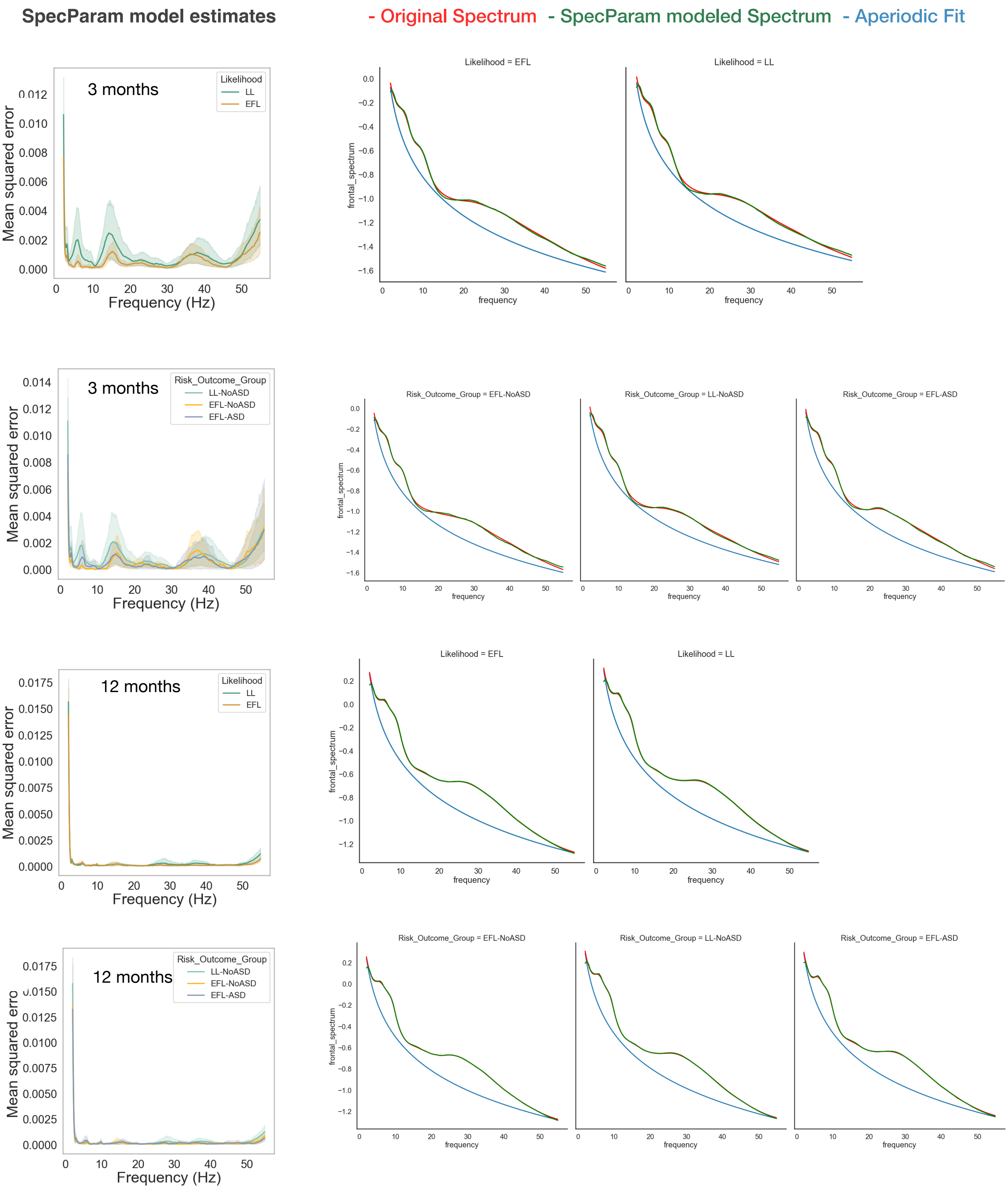
