## Supplemental Tables for "Changes in early aperiodic EEG activity are linked to autism diagnosis and language development in infants with family history of autism"

Supplemental Table 1: Infant Sibling Project Summary of enrollment and EEG collection and exclusion.

|  | Infant Sibling Project.  Total Enrolled Participants = 233  Low Likelihood (LL) = 98*  Elevated Familial Likelihood (EFL) = 132 | | | | |
| --- | --- | --- | --- | --- | --- |
| **Participant EEG Data Retention** | | | | | |
|  | Likelihood | Visit Occurred | EEG Collected | EEG Excluded Due to Behavior** | EEG met quality  (% of Visits) |
| 3 m | LL | 18 | 18 | 16 | 14 (77.8%) |
|  | EFL | 41 | 40 | 34 | 32 (78.0%) |
| 12 m | LL | 80 | 77 | 72 | 71 (88.8%) |
|  | EFL | 107 | 98 | 87 | 84 (78.5%) |

*3 LL participants with later ASD diagnoses removed.

** Behavioral reasons for exclusion include participant sleeping or crying throughout recording

|  | Infant Screening Project  Total Enrolled Participants = 194  Low Likelihood (LL) = 112*  Elevated Familial Likelihood (EFL) = 82 | | | | |
| --- | --- | --- | --- | --- | --- |
| **Participant EEG Data Retention** | | | | | |
|  | Likelihood | Visit Occurred | EEG Collected | EEG Excluded Due to Behavior** | EEG met quality  (% of Visits) |
| 3 m | LL | 50 | 49 | 48 | 45 (90.0%) |
|  | EFL | 32 | 32 | 32 | 25 (78.1%) |
| 12 m | LL | 104 | 97 | 96 | 86 (82.7%) |
|  | EFL | 70 | 65 | 65 | 60 (86.0%) |

Supplemental Table 2: Infant Screening Project Summary of enrollment and EEG collection and exclusion.

*4 LL participants with later ASD diagnoses removed.

** Behavioral reasons for exclusion include participant sleeping or crying throughout recording

|  | Likelihood | Total | N = Infant Sibling/Infant Screening | |
| --- | --- | --- | --- | --- |
|  |  |  | 64-channel net | 128-channel net |
| 3 m | LL | 59 | 1/0 | 13/45 |
|  | EFL | 57 | 6/0 | 26/25 |
| 12 m | LL | 157 | 12/0 | 59/86 |
|  | EFL | 144 | 27/0 | 57/60 |

|  |  |  | N = Infant Sibling/Infant Screening | | | |
| --- | --- | --- | --- | --- | --- | --- |
|  | Likelihood | Total | 64-channel net | | 128-channel net | |
|  |  |  | 200 Amp | 300 Amp | 200 Amp | 300 Amp |
| 3 m | LL | 59 | 1/0 | 0/0 | 5/0 | 8/45 |
|  | EFL | 55 | 5/0 | 0/0 | 1/0 | 24/25 |
| 12 m | LL | 156 | 12/0 | 0/0 | 10/0 | 46/86 |
|  | EFL | 121 | 26/0 | 0/0 | 17/0 | 34/60 |

Supplemental Table 3: Summary of EEGs analyzed in paper using 64 and 128 channel nets.

Supplemental Table 4: Summary of EEGs analyzed in paper using NetAm 200 vs NetAmp 300 EGI amplifiers.

Note - amplifier information was not available for all participants in the Infant Sibling Study.

**Supplemental Table 5:** To assess whether differences in NetAmp200 and NetAmp300 affected EEG data quality, we selected a subset of participants matched on the following criteria: (1) net type (128-channel net), (2) sampling rate (250 Hz), and (3) without an autism diagnosis from the Infant Siblings Project. This resulted in 22 infants with NetAmp 200 and 65 infants with NetAmp 300. Thus, we randomly selected 22 infants from NetAmp 300, matching the sample size of those collected with the NetAmp 200.

|  | Measure | NetAmp 200 (Mean(SD)) | NetAmp 300 (Mean(SD)) | Test Statistic | p-value |
| --- | --- | --- | --- | --- | --- |
| Data Quality Metrics | Signal-to-Noise ratio | -0.38 (7.19) | -1.81(7.14) | Mann-Whitney U =0.66 | p= 0.51 |
|  | Percent ICs Rejected | 36 (9) | 40 (12) | t-test=-1.14 | p=0.26 |
|  | Percent Good Channels | 91 (6) | 91 (5) | Mann-Whitney U =243 | p=0.99 |

**Supplemental Table 6:** To assess whether differences in EEG acquisition paradigms between Infant Sibling Project (Study 1) and Infant Screening Project (Study 2) affected data quality and EEG measures of interest, we selected a subset of participants matched on the following criteria: (1) age range (360–400 days), (2) net type (128-channel net), (3) amplifier model (NetAMP 300), (4) sampling rate (500 Hz), and (5) without an autism diagnosis. This resulted in 24 infants from Study 1 and 64 infants from Study 2. Thus, we randomly selected 24 infants from Study 2, matching the sample size of Study 1. Statistical comparisons were conducted based on the distribution of the measure of interest. Note that we do not find evidence for EEG data quality metrics nor our main measures of interests between Study 1 and Study 2.

|  | Measure | Study 1  (Mean(SD)) | Study 2  (Mean(SD)) | Test Statistic | p-value |
| --- | --- | --- | --- | --- | --- |
| Data Quality Metrics | Signal-to-Noise ratio | -0.45 (5.31) | -2.47(7.12) | Mann-Whitney U =338 | p= 0.31 |
|  | Percent ICs Rejected | 0.39 (0.1) | 0.43 (0.1) | t-test= -1.60 | p=0.12 |
|  | Percent Good Channels | 93.86 (3.31) | 92.73 (4.75) | Mann-Whitney U = 297.5 | p=0.65 |
|  | Percent Good Segments | 70.17 (7.39) | 71.25 (4.33) | Mann-Whitney U = 257 | p=0.69 |
| Main EEG measures of Interest | Aperiodic Offset | 0.2 (0.20) | 0.2 (0.18) | t-test= 0.04 | p=0.97 |
|  | Aperiodic Exponent | 1.10 (0.1) | 1.09 (0.09) | t-test= 0.55 | p=0.58 |
|  | Aperiodic Total Power | -45.60 (8.14) | -44.92 (9.02) | t-test= -0.27 | p=0.79 |
|  | Periodic Total Power | 7.60 (2.69) | 7.21 (2.71) | t-test= 0.50 | p=0.62 |

**Supplemental Table 7:** 3-month sensitivity analysis of EEG feature differences between EFL and LL Groups (EFL first)

All participants (EFL = 57, LL = 59); 64 channel nets removed (EFL = 51; LL = 58); 200Amp removed (EFL=51; LL = 53)

|  |  | Likelihood | | Statistics | |
| --- | --- | --- | --- | --- | --- |
| EEG feature, Mean (SD) | Sensitivity Analysis | EFL | LL | Mann-Whitney U Statistic | Mann-Whitney p-value |
| **Absolute Gamma** | | | | | |
|  | All participants | -19.15 (3.22) | -18.05 (2.68) | 1268 | 0.023 |
|  | 64 channel nets removed | -19.33 (2.85) | -18.08 (2.69) | 1112 | 0.026 |
|  | 200 Amp EEGs removed | -19.21 (2.83) | -18.22 (2.66) | 1083 | 0.081 |
| **Aperiodic Offset** | | | | | |
|  | All participants | -0.18 (0.19) | -0.13 (0.15) | 1486 | 0.282 |
|  | 64 channel nets removed | -0.17 (0.18) | -0.13 (0.16) | 1359 | 0.468 |
|  | 200 Amp EEGs removed | -0.16 (0.18) | -0.14 (0.16) | 1283 | 0.658 |
| **Aperiodic Slope** | | | | | |
|  | All participants | 1.07 (0.14) | 1.03 (0.11) | 1968 | 0.114 |
|  | 64 channel nets removed | 1.09 (0.12) | 1.03 (0.11) | 1824 | 0.036 |
|  | 200 Amp EEGs removed | 1.09 (0.12) | 1.03 (0.11) | 1657 | 0.047 |
| **Total Aperiodic Activity** | | | | | |
|  | All participants | -63.83 (10.35) | -59.54 (8.51) | 1187 | 0.006 |
|  | 64 channel nets removed | -64.25 (9.17) | -59.63 (8.56) | 1059 | 0.011 |
|  | 200 Amp EEGs removed | -63.86 (9.00) | -59.95 (8.40) | 1007 | 0.025 |
| **Periodic Gamma** | | | | | |
|  | All participants | 2.19 (0.85) | 1.96 (1.13) | 1989 | 0.090 |
|  | 64 channel nets removed | 2.18 (0.85) | 1.96 (1.14) | 1739 | 0.115 |
|  | 200 Amp EEGs removed | 2.19 (0.85) | 1.91 (1.16) | 1663 | 0.043 |

Supplemental Table 8: 3-month comparison of EEG feature differences Outcome Groups

| 3-month EEG data | Outcome | | | Statistics | |
| --- | --- | --- | --- | --- | --- |
| EEG feature, Mean (SD) | LL-noASD | EFL-NoASD | EFL-ASD | Kruskal Wallis  H statistic | Kruskal Wallis  p-value |
| Absolute Gamma | -18.10 (2.86) | -18.93 (4.03) | -18.69 (2.51) | 2.10 | 0.351 |
| Aperiodic Offset | -0.14 (0.16) | -0.19 (0.18) | -0.14 (0.18) | 1.13 | 0.534 |
| Aperiodic Slope | 1.03 (0.11) | 1.05 (0.14) | 1.08 (0.10) | 2.23 | 0.328 |
| Total Aperiodic Activity | -59.68 (9.39) | -63.32 (12.01) | -62.34 (8.98) | 4.12 | 0.127 |
| Periodic Gamma | 1.94 (1.13) | 2.21 (0.98) | 2.26 (0.56) | 4.26 | 0.119 |

**Supplemental Table 9**

|  | **Aperiodic Total Power** | | | | **Aperiodic Offset** | | | | **Aperiodic Exponent** | | | | **Absolute Gamma Power** | | | |
| --- | --- | --- | --- | --- | --- | --- | --- | --- | --- | --- | --- | --- | --- | --- | --- | --- |
| *Predictors* | *std. Beta* | *std. Error* | *Statistic* | *p* | *std. Beta* | *std. Error* | *Statistic* | *p* | *std. Beta* | *std. Error* | *Statistic* | *p* | *std. Beta* | *std. Error* | *Statistic* | *p* |
| (Intercept) | -0.83 | 0.20 | -27.64 | **<0.01** | -0.80 | 0.19 | -3.86 | **<0.01** | -0.12 | 0.28 | 36.81 | **<0.01** | -0.76 | 0.21 | -25.68 | **<0.01** |
| Age | 1.84 | 0.24 | 7.66 | **<0.01** | 1.92 | 0.22 | 8.67 | **<0.01** | 0.66 | 0.32 | 2.07 | **0.04** | 1.66 | 0.23 | 7.30 | **<0.01** |
| Outcome  [LL-NoASD] | 0.33 | 0.21 | 1.57 | 0.12 | 0.27 | 0.20 | 1.36 | 0.18 | -0.06 | 0.29 | -0.21 | 0.84 | 0.26 | 0.21 | 1.20 | 0.23 |
| Outcome [EFL-NoASD] | -0.10 | 0.25 | -0.39 | 0.70 | -0.05 | 0.24 | -0.20 | 0.84 | 0.10 | 0.35 | 0.28 | 0.78 | -0.06 | 0.26 | -0.24 | 0.81 |
| sex | -0.04 | 0.13 | -0.35 | 0.73 | -0.10 | 0.12 | -0.84 | 0.40 | -0.15 | 0.18 | -0.84 | 0.40 | -0.11 | 0.14 | -0.78 | 0.44 |
| age days | 0.11 | 0.06 | 1.91 | 0.06 | 0.16 | 0.06 | 2.84 | **0.01** | 0.13 | 0.08 | 1.61 | 0.11 | 0.13 | 0.06 | 2.26 | **0.03** |
| Age × Outcome [LL-NoASD] | -0.65 | 0.28 | -2.29 | **0.02** | -0.67 | 0.26 | -2.56 | **0.01** | -0.21 | 0.38 | -0.55 | 0.59 | -0.44 | 0.27 | -1.65 | 0.10 |
| Age × Outcome [EFL-NoASD] | -0.39 | 0.34 | -1.15 | 0.25 | -0.54 | 0.31 | -1.72 | 0.09 | -0.45 | 0.45 | -1.01 | 0.32 | -0.13 | 0.32 | -0.42 | 0.68 |
| **Random Effects** | | | | | | | | | | | | | | | | |
| σ^2^ | 58.79 | | | | 0.02 | | | | 0.01 | | | | 5.09 | | | |
| τ_00_ | 5.77 _StudyID_ | | | | 0.00 _StudyID_ | | | | 0.00 _StudyID_ | | | | 1.37 _StudyID_ | | | |
| ICC | 0.09 | | | | 0.14 | | | | 0.15 | | | | 0.21 | | | |
| N | 73 _StudyID_ | | | | 73 _StudyID_ | | | | 73 _StudyID_ | | | | 73 _StudyID_ | | | |
| Observations | 146 | | | | 146 | | | | 146 | | | | 146 | | | |
| Marginal R^2^ / Conditional R^2^ | 0.513 / 0.556 | | | | 0.556 / 0.618 | | | | 0.076 / 0.218 | | | | 0.495 / 0.602 | | | |
